# A Systematic Review and Meta Synthesis Protocol for Examining the Factors behind African Nurses’ & Midwives Migration

**DOI:** 10.1101/2023.02.27.23286495

**Authors:** Stephanopoulos Kofi Junior Osei, Michael Owusu Barfi, Sandra Frimpong, Dorinda Dela Bosro, Christopher Asamoah Fosu, Daniel Owusu, Deborah Ntriwaa Amoako-Mensah, Sammy Kwantwi Barimah, Jerry Kofi Esinu Agbavor, Bertha Awo Agbesi Delanyo

## Abstract

**Background:** There has been an increased rise of African nurses and midwives migrating to high income countries despite varying policies and restrictions to promote retention. The need to comprehensively understand the factors influencing the exodus is critical for policy formulation at the global, national, and facility levels.

**Aim:** To provide a comprehensive and a systematic review of the factors influencing African nurses and midwives’ migration to high income countries and the factors influencing such migrations.

**Methods:** A systematic review and meta-synthesis design guided by the Preferred Reporting Items for Systematic Reviews and Meta-Analyses (PRISMA) Statement guidelines would be used. Databases including CINAHL, Pubmed, MEDLINE, and EbscoHost will be searched using a PICOS selection criteria framework. Two independent reviewers would be involved in data extraction and meta-synthesis. A third reviewer would provide an arbitrary judgement when disagreements persist after discussion. A meta-synthesis and thematic analysis would be utilised to identify emerging themes and reporting themes identified in the literature.

**Ethics and dissemination:** The study does not require ethical approval. The findings would be published in peer-review journals and presented at conferences.

**PROSPERO registration number:** CRD42023395013

**Strengths and limitations:**

- The study would shed a comprehensive light on factors influencing the migration of African trained nurses outside the continent. This is relevant in informing policies that promote retention.
- The review is conducted following the guidelines of Preferred Reporting Items for Systematic Reviews and Meta-Analyses statement (PRISMA). The methodology is rigorous and meets the standards of the PRISMA guidelines.
- Studies published in any language other than English would be excluded.
- The period restriction set could also limit the number of studies found for a comprehensive review.

## Background

Nursing migration from low- and middle-income countries (LMICs) in Africa to more developed nations has become an increasingly ubiquitous phenomenon.^1^ According to the Organisation for Economic Co-operation and Development (OECD)2, LMICs, especially Ghana, Nigeria, Kenya, and Zimbabwe, have become relatively prominent contributors of foreign-trained nurses to OECD countries, which are mostly high-income countries.

The migration of healthcare professionals, particularly nurses, from Africa to developed countries is a significant issue that has far-reaching consequences for both the sending and receiving countries. As it stands, the global distribution of nurses is highly unequal, with Africa having the highest shortage and the worst ratio of nurses to patients.^3^ This imbalance is further exacerbated by the migration of African nurses to more developed countries, which results in domestic shortages of nurses and worsens the nurse-patient ratio.^4^ Although the migration of African nurses mostly favours the receiving countries, there are certainly ethical implications and the issue of offsetting the standards of practice in these countries.^5^

There are a myriad of policies and restrictions established by both national institutions and global organisations to regulate the influx of African nurses to developed nations. Of such policies is the notable WHO ‘red list” which bans active recruitment of nurses and other healthcare professionals from 47 countries with most pressing healthcare workforce challenges. About 70% of the listed nations are countries from the African region.^6^ As indicated earlier, different policies are being instituted at national level. Examples include Namibia incentives (subsidised cars and home ownership) for nursing students and registered nurses, bonding methods by countries like Ghana, South Africa, Zimbabwe and Lesotho which require nurse graduates to repay their training cost to their government before they can migrate.^7^

However, it appears that these measures have not been entirely effective in mitigating the nurses’ exodus in droves. Between 2021 and 2022, countries like Ghana, Nigeria, and Nepal, which are all on the WHO ban list, contributed 19% of nurses and midwives recruited to the UK.^8^ Additionally, governments of Ghana and Kenya are partnering with the UK and other countries in the form of memorandums that support nursing recruitments.^9^ Although these memorandums provide financial benefits to the sending countries, perhaps it is informed by the relentless determination of nurses and midwives to migrate regardless of how disincentive the process has become.

Previous research has investigated the factors that contribute to the migration of nurses and midwives, with studies such as Nair et al.^10^ and Dywilli et al.^11^ providing valuable insights. However, there is a notable lack of systematic reviews specifically focusing on the African context, and the existing studies in this area are somewhat outdated.^12^ This highlights the need for a comprehensive, up-to-date systematic review of the literature on the migration of nurses and midwives in Africa

### Objective

The current systematic review is being conducted to provide a comprehensive and systematic review of the factors behind their relentless determination of African nurses and midwives to migrate to high income countries. The goal is to contribute to the growing body of evidence that could be used to inform policy making regarding the retention of African nurses and midwives.

## Method

### Search Strategy

We will employ a systematic review and meta-synthesis design. The review would follow Preferred Reporting Items for Systematic Reviews and Meta-Analyses (PRISMA) 2020 updated guidelines.^13^ Published articles between 2010 to March, 2023 would be searched on databases including PubMed and CINAHL on EbscoHost. Grey literature using selected keywords would be conducted on google. The search terms would include the keywords for the concept being examined, text words (synonymous to the concept-related keywords), and medical Subject Headings (MeSH). Search terms such as “nurses”, “midwives”, “migration”, “emigration”, “brain drain”, “push factors”, “pull factors” and will be combined using Boolean operators to generate a comprehensive search strategy (Table 1). The search strategy would be modified to suit the operations of the two databases. A search strategy to be conducted on PubMed has been provided as an example and is available in supplemental appendix 1. Additionally, reference lists of identified studies would be hand-searched to identify relevant studies. All records identified would be Rayyan (https://rayyan.ai/) – a web app for article screening and filtering. ^14^

**Table 1.**
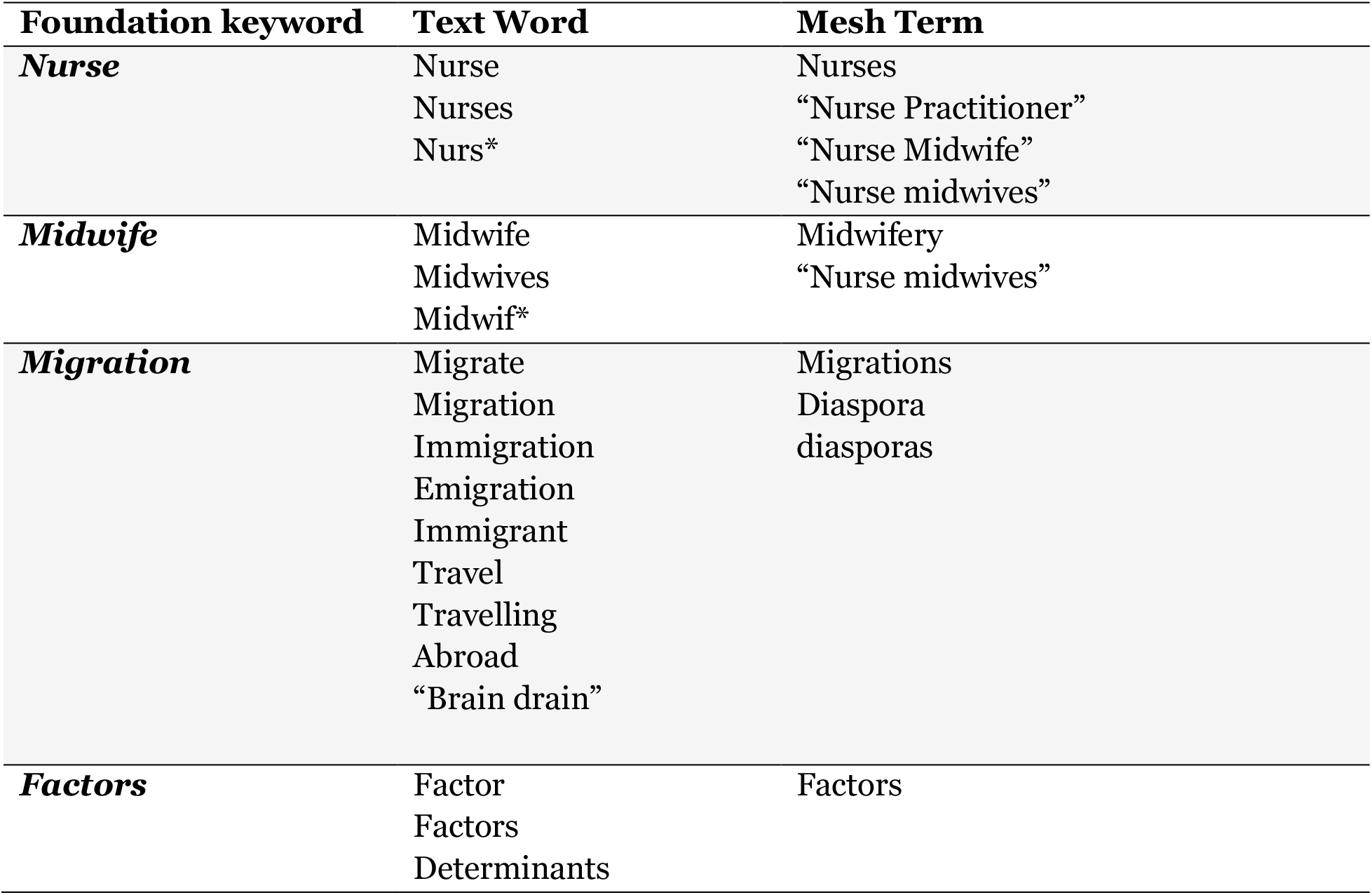
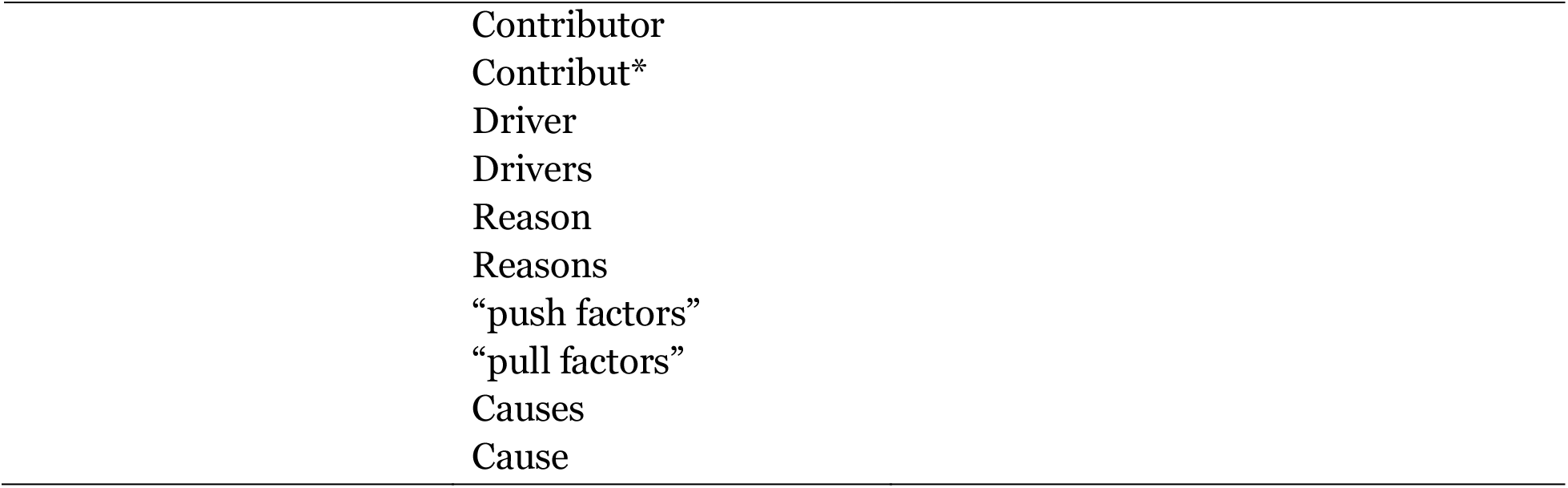
Search Terms to be used in the Review

### Selection Criteria

The PICOS framework (population, issues, context, outcomes, and study design) (Table 2) would be used as selection criteria.^15^ Essentially original qualitative or quantitative and mixed method studies on factors influencing African nurses’ and midwives’ migration outside Africa would be considered during the review. These studies would provide comprehensive insights on the phenomenon. Papers would be excluded if they were not published in English (due to a lack of translators).

**Table 2.**
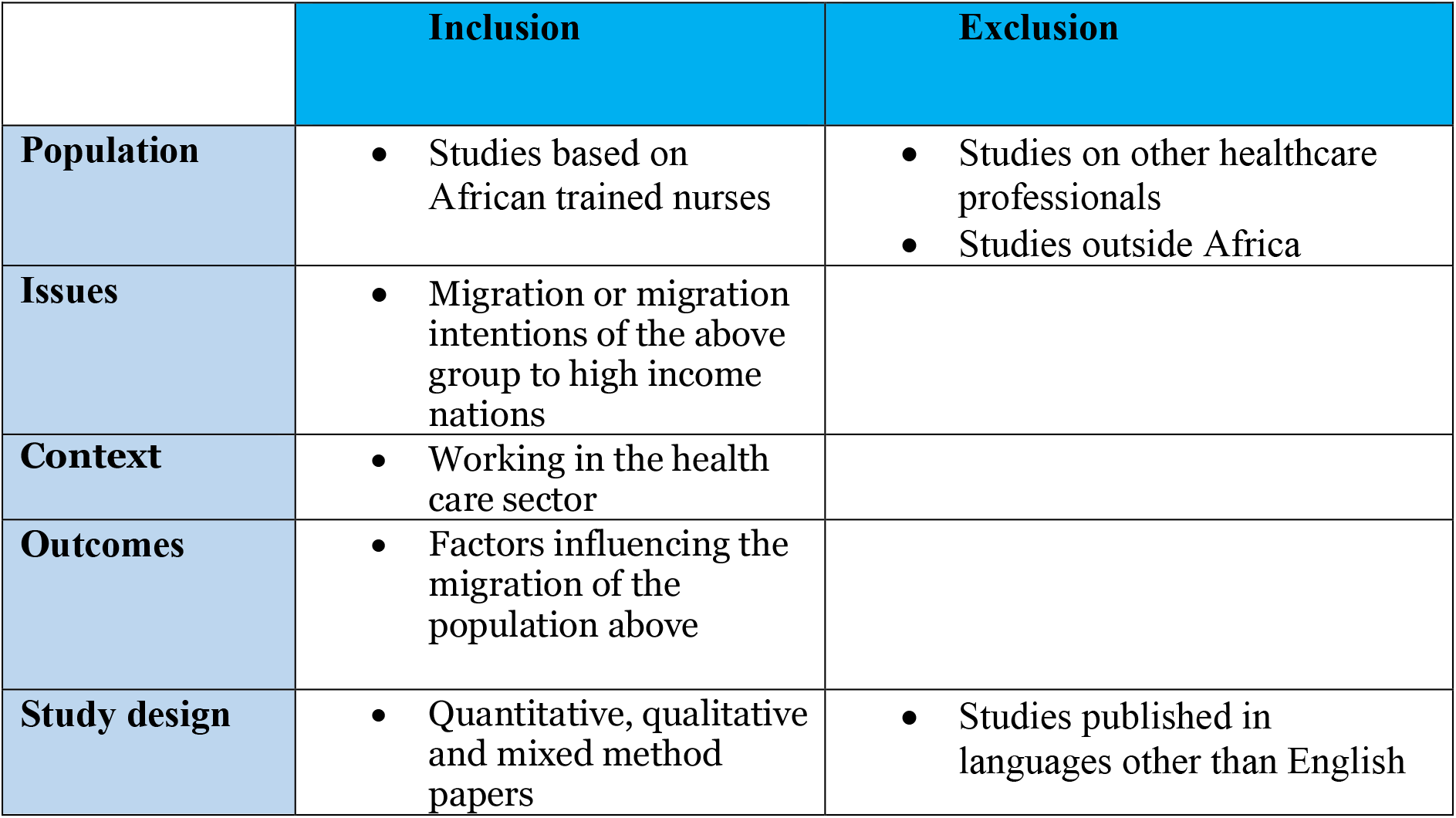
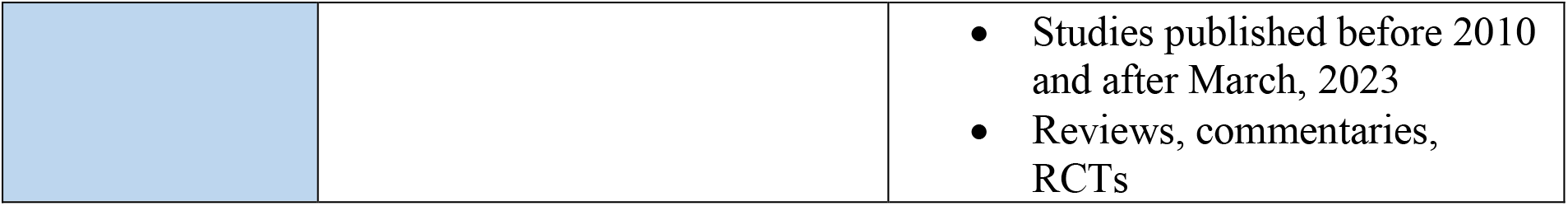
The Population, Issues, Context, Outcomes and Study design (PICOS)

|  | Inclusion | Exclusion |
| --- | --- | --- |
| **Population** | Studies based on African trained nurses | Studies on other healthcare professionals<br>Studies outside Africa |
| **Issues** | Migration or migration intentions of the above group to high income nations |  |
| **Context** | Working in the health care sector |  |
| **Outcomes** | Factors influencing the migration of the population above |  |
| **Study design** | Quantitative, qualitative and mixed method papers | Studies published in languages other than English |
|  |  | • Studies published before 2010 and after March, 2023<br>• Reviews, commentaries, RCTs |

### Selection method

All records would be screened for duplicate studies and all duplicates would be deleted using Rayyan. Four pairs of independent reviewers (MBO and SF, DDB and CFA, DO and DNA, SKB and JKEA) would screen 25% of the total number of titles and abstracts each to identify studies that are potentially related to African nurse and midwife migration (Fig 1). We would then obtain the full text of these articles, which would be screened to identify if they fit the inclusion criteria. Conflicts in selection would be resolved through discussions and if arbitrary decision by SKJO.

**Fig 1.**
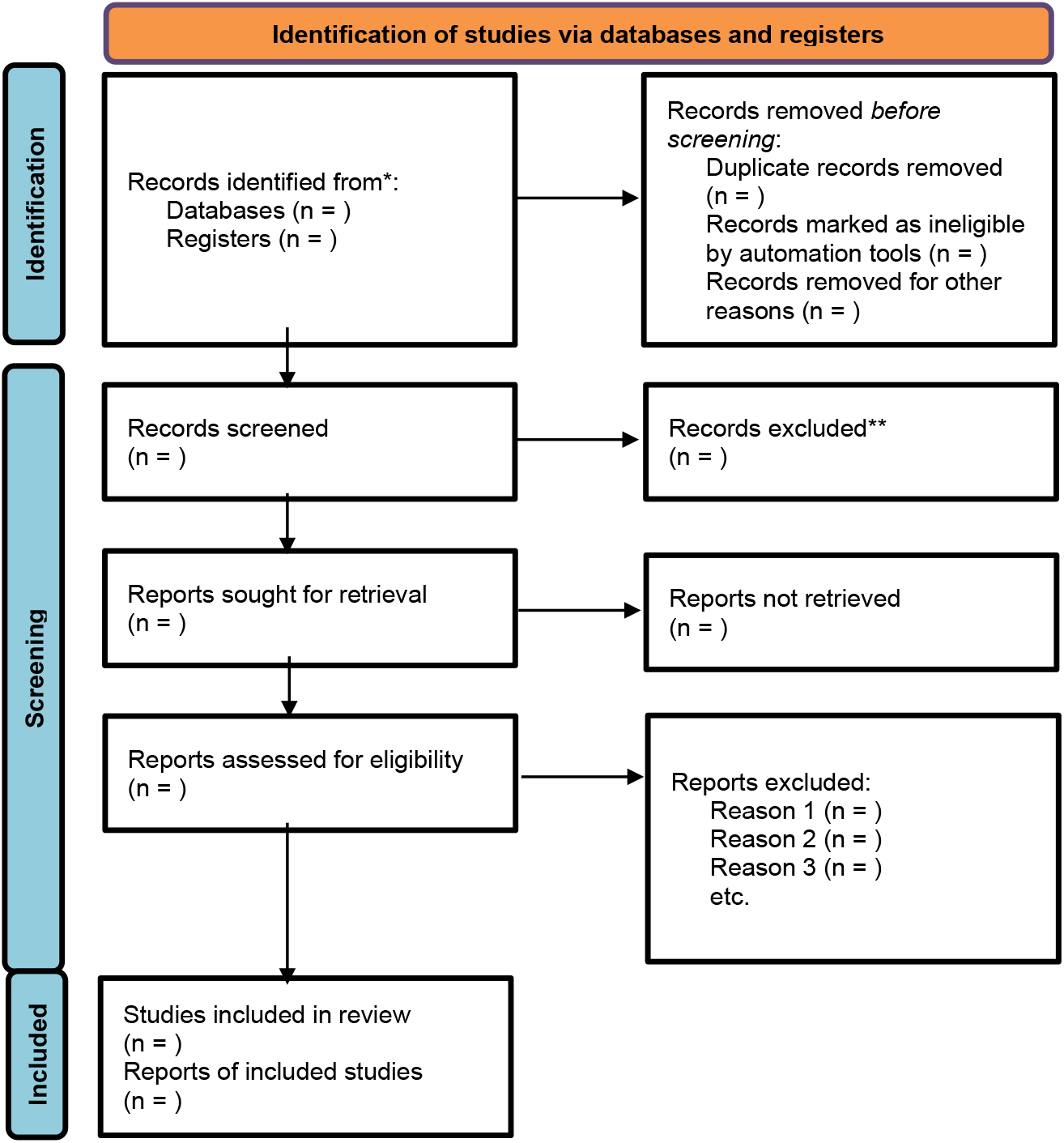
Prisma Flow Diagram for Systematic Review.

**Fig 2.**
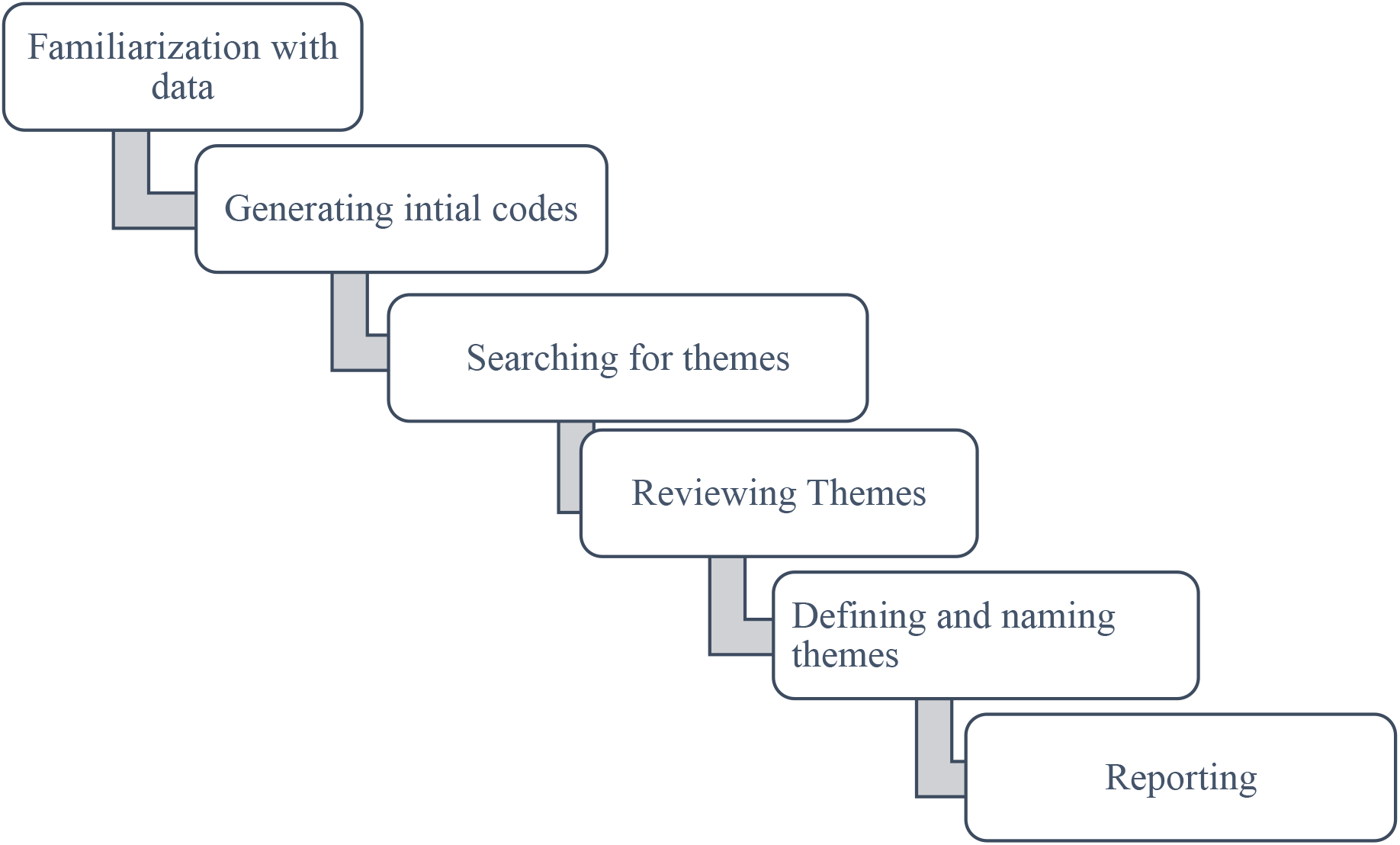
Thematic analysis: Braun & Clarke Six interactive phases.

### Quality assessment

A pair of independent reviewers (SKJO and BDAA) would appraise identified studies using the Mixed Method Appraisal Tools (MMAT) developed by Pluye et al.^16^ The tools can be used to appraise the methodological quality and assessing risk of bias of empirical studies. MMAT is appropriate for scoring studies using either qualitative approach, quantitative approach or mixed. However, studies would not be excluded due to their appraisal scores as to provide a comprehensive picture on determinants of nurse migration in Africa. The scores of these independent reviews would be discussed and conflicts would be resolved with the involvement of a third reviewer (MBO).

### Data analysis and synthesis

Thematic analysis would be guided by Braun & Clarke ^17^ six phases of thematic analysis. Two independent authors would familiarise themselves with studies identified by repeated reading and then an active reading of the results/findings of studies appraised to identify meanings and patterns. Initial open codes would be produced from the data and codes would be managed using Microsoft Excel 2016. Codes would then be compared and organised into subcategories and broader categories to provide themes. These would be revised and refined (collapsed into each other or broken down). The emerging themes would be discussed by the entire team until a final consensus is reached.

### Ethics and dissemination

Ethics approval is not applicable to this study as data would be gathered from already published studies. We intend to disseminate the findings through peer-reviewed journals and conference presentations. Protocol amendments would be conducted in the registry (PROSPERO) should there be any.

## Discussion

In this protocol, we have provided the background information and design of a systematic review and meta-synthesis of original researchers examining the factors influencing migration of African nurses and midwives outside the continent. The review is projected to identify the push and pull factors impacting the high levels of migration. The results are intended to inform arising global, national and facility policies to improve the retention of nurses and midwives in Africa.

The study findings would be reported in concordance with the guidelines from Preferred Reporting Items for Systematic Reviews and Meta-Analysis (PRISMA) statement.

A major limitation of this review is the exclusion of studies published in languages other than English. This excluding mean we may miss relevant studies publish in other languages especially French and Portuguese which are all major national languages in many African countries and other nations outside the continent.

## Conclusion

The systematic review on factors influencing the migration of nurses and midwives trained in African nations to high income country is vital in understanding a comprehensive picture of the phenomenon and supporting policies and strategies promoting retention.

## Author contributions

SKJO was responsible for formulating the idea of the study. SKJO, MBO, DO, SF, DNA were responsible for literature review, and identifying research gaps. SKJO was responsible for the study design. SKB, JKEA, BDAA, DDB, SKJO, MBO, DO, SF, DNA, CFA would review evidence and are responsible for report writing.

## Data Availability

All data produced in the work would be contained in the manuscript

## Appendix 1.

### Search Strategy for Pubmed

((nurse[tiab] OR nurses[tiab] OR “nurse practioner”[tiab]) OR (midwives[tiab] OR midwife[tiab] OR “nurse midwife”[tiab] OR nurse-midwife[tiab]) AND (migrate[tiab] OR migration[tiab] OR migra*[tiab] OR immigration[tiab] OR emmigration[tiab] OR immigrate[tiab] OR emmigrate[tiab]) AND (factors[tiab] OR determinants[tiab] OR causes [tiab])

